# Discrete SIR modelling using empirical infection data shows that SARS-CoV-2 infection provides short-term immunity

**DOI:** 10.1101/2020.06.03.20120113

**Authors:** Andrew McMahon, Nicole C. Robb

## Abstract

**Background:** The novel coronavirus SARS-CoV-2, which causes the COVID-19 disease, has resulted in a global pandemic. Since its emergence in December 2019, the virus has infected millions of people, caused the deaths of hundreds of thousands and resulted in incalculable social and economic damage. Understanding the infectivity and transmission dynamics of the virus is essential for understanding how best to reduce mortality whilst ensuring minimal social restrictions to the lives of the general population. Anecdotal evidence is available, but detailed studies have not yet revealed whether infection with the virus results in immunity.

**Objective:** The objective of the study was to use mathematical modelling to investigate the reinfection frequency of COVID-19.

**Methods:** We have used the SIR (Susceptible, Infected, Recovered) framework and random processing based on empirical SARS-CoV-2 infection and fatality data from different regions to calculate the number of reinfections that would be expected to occur if no immunity to the disease occurred.

**Results:** Our model predicts that cases of reinfection should have been observed by now if primary SARS-CoV-2 infection did not protect from subsequent exposure in the short term, however, no such cases have been documented.

**Conclusions:** This work concludes that infection with the SARS-CoV-2 virus provides short-term immunity to reinfection and therefore provides a useful insight for serological testing strategies, lockdown easing and vaccine design.

## Introduction

The novel coronavirus SARS-CoV-2 is thought to have originated in China in late 2019, and has since spread globally, resulting in the COVID-19 pandemic. In the eight months since the first confirmed case, the virus has resulted in 24 million confirmed infections, over 820,000 deaths, and caused huge social and economic damage.

SIR (Susceptible, Infected, Recovered) modelling uses a set of differential equations to determine how the number of infected and recovered individuals changes over time given a specified rate of infection and recovery. It was first used in 1927 by Kermack and McKendrick [1] and has since been used to model epidemics from Acquired Immune Deficiency Syndrome (AIDS) [2] to SARS [3]. Variations of SIR modelling have been used during the COVID-19 pandemic to look at the varying burden on healthcare systems based on public health intervention [4], the absence of a stable disease-free equilibrium [5] and infection rate [6], as well as the eventual size of the overall pandemic [7]. An extension of the model has also been used to simulate the changing death rate as a function of the number of individuals infected, and it was found that an equilibrium point was reached where there are no further reinfections [8].

In this study, we used an extension to the SIR framework which distinguished between infected and reinfected individuals to model empirical data taken from a compiled COVID-19 dataset [9], in order to investigate the reinfection frequency of the disease. The results of this analysis showed that a small number of cases classified as ‘reinfections’ should have occurred; however, no definitive cases of reinfection have been reported in the scientific literature to date. This suggests that primary SARS-CoV-2 infection is effective at preventing reinfection in the short term.

## Methods

### Data sources

We used national infection and mortality data from a variety of sources to investigate the reinfection dynamics of SARS-CoV-2. Unless specified, the national data on infections and deaths from SARS-CoV-2 was acquired from the ‘Our World in Data’ database compiled by the Oxford Martin School at the University of Oxford [9]; the hospitalisation cases in Switzerland were obtained from the Federal Office of Public Health in Switzerland [10]; the data for the city of New York was obtained from the New York City Health website [11]; the population of New York City was obtained from the 2019 New York census [12] and the recovery data for Germany was sourced from Trading Economics [13] which gets data from WHO (World Health Organisation). For each geographical region, the data was taken from the date of the first recorded infection up until the 17^th^ May 2020 when the data was accessed.

### Choice of geographical regions

The simulations were initially completed for the United Kingdom (UK) where, at the time the data was accessed, there were a high number of confirmed cases. Australia was selected as an example of a region with low numbers of recorded cases in order to investigate the limit of expected reinfections; Germany was selected as it was one of few countries with recorded recovery data; Italy was studied as the number of infections and deaths had peaked by the 17^th^ May 2020; Singapore was unique as a city-state so population density for the nation was very high; Switzerland was selected as hospitalisation data was available at the time the data was accessed, and the United States of America (USA) as a whole was compared with New York City, which was the worst affected state in the USA at the time.

### Assumptions

A number of assumptions have been made. Where possible, they have been made so that the number of reinfections is underestimated. These assumptions are:

1. There is a large lag time for recovery to take place (28 days) [14,15].
2. The incubation period was modelled as 6 days [16].
3. The model does not consider social distancing or shielding and so assigns an equal probability of an infection to all individuals.
4. Not all infections have been recorded due to lack of testing, misdiagnosis or asymptomatic infection. [17]
5. Infections and recoveries are not necessarily recorded on the date that they first occurred.
6. There is no emigration out of, or immigration into, a population of interest.
7. The model assumes a homogeneous population density, with no societal structure (e.g. equal residents per household).

### The model

We based our model on the compartmental SIR framework, but differentiated between initial and subsequent infections, resulting in a six-state model (Susceptible, Infected, Recovered, Infected (two or more times), Recovered (two or more times) and Deceased) (Figure 1 & Table 1). The number of infections and deaths each day were taken from national statistics (as described above). Where available, recovery data was used, and otherwise recoveries were modelled with a 28 day lag time (with the number of recoveries representing those individuals that did not die during the 28 day recovery time). ‘Recovered’ individuals were selected stochastically from the populations of the states 28 days prior [14,15]; ‘infected’ individuals from the populations of the states 6 days prior (due to the incubation period [16]), and ‘deceased’ individuals from the populations of the states 1 day prior.

**Figure 1.**
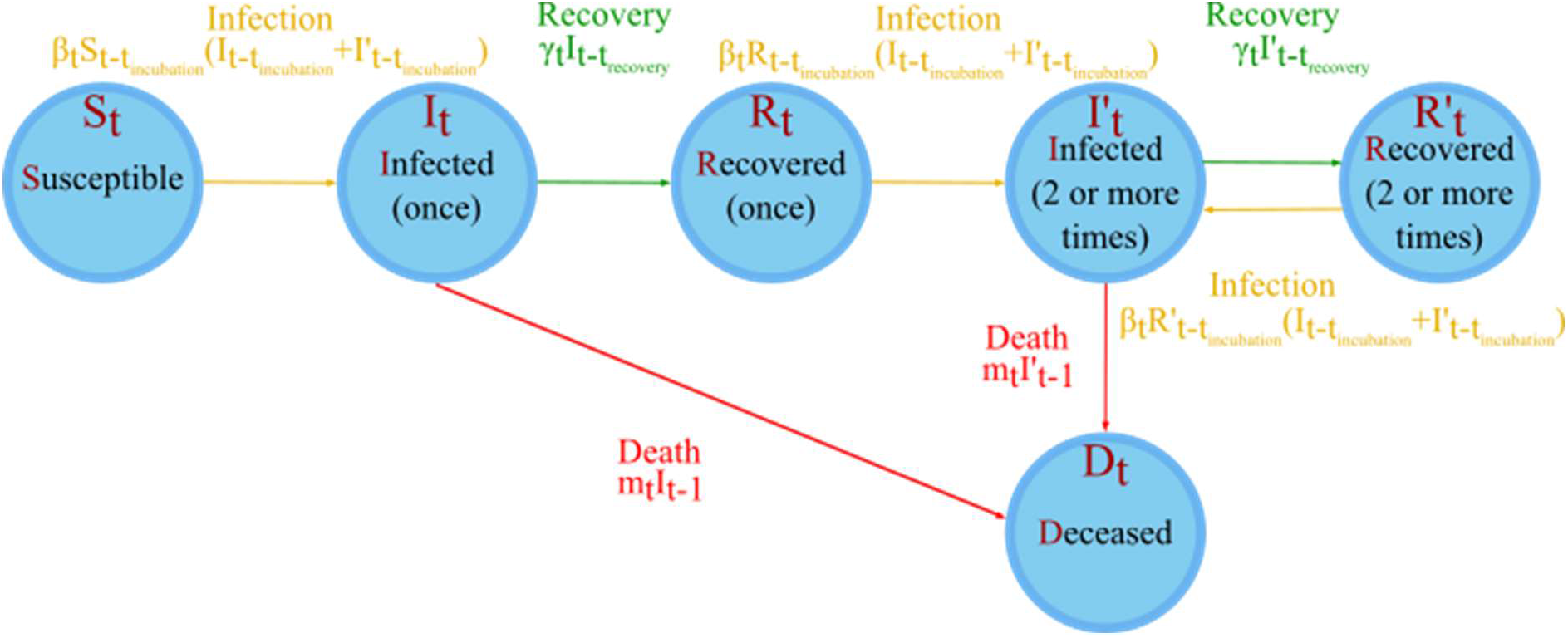
A simple representation of the model. S_t_ represents the number of persons susceptible to infection who have had no prior infections on day t; I_t_ is the number of people currently infected for the first time on day t; R_t_ is the number of people who have recovered once on day t; I’_t_ is the number of people who have been infected two or more times and are infected on day t; R ’t is the number of people who have recovered two or more times and are not infected on day t of the model and D_t_ is the number of deceased persons on day t of the model. Further symbols are defined in Table 1.

**Table 1.** Definition of parameters in the model.

|  |  |
| --- | --- |
| $x_i$ | Random number |
| $t$ | The number of days into the simulation. $t=1$ for the day of the first infection |
| $t_{\text{recovery}}$ | The average number of days for recovery. Here, we have used $t_{\text{recovery}}=28$ |
| $t_{\text{incubation}}$ | The average number of days for an infection to be seen. Here, we have used $t_{\text{incubation}} = 6$ |
| $t_{\text{max}}$ | The number of days over which the simulation is run |
| $S_t$ | The number of susceptible on day $t$ |
| $I_t$ | The number of Infected (once) on day $t$ |
| $R_t$ | The number of Recovered (once) on day $t$ |
| $I'_t$ | The number of Infected (two or more times) on day $t$ |
| $R'_t$ | The number of Recovered (two or more times) on day $t$ |
| $D_t$ | The number of deceased from SARS-CoV-2 on day $t$ |
| $B_t$ | The infection rate on day $t$ |
| $\gamma$ | The recovery rate on day $t$ |
| $m_t$ | The death rate on day $t$ |
| $N$ | The total population in the model<br>$N = S_t + I_t + R_t + I'_t + R'_t + D_t \quad \forall t$ |
| $N_t^{\text{infected}}$ | The number of infected individuals on day $t$<br>$N_t^{\text{infected}} = I_t + I'_t$ |
| $N_t^{\text{uninfected}}$ | The number of uninfected individuals on day $t$<br>$N_t^{\text{uninfected}} = S_t + R_t + R'_t$ |
| $n_t^{\text{infected}}$ | The number of new infections on day $t$<br>$n_t^{\text{infected}} = \beta_t S_{t-t_{\text{incubation}}} (I_{t-t_{\text{incubation}}} + I'_{t-t_{\text{incubation}}}) + \beta_t R_{t-t_{\text{incubation}}} (I_{t-t_{\text{incubation}}} + I'_{t-t_{\text{incubation}}}) + \beta_t R'_{t-t_{\text{incubation}}} (I_{t-t_{\text{incubation}}} + I'_{t-t_{\text{incubation}}})$ |
| $n_t^{\text{recovered}}$ | The number of recovering individuals on day $t$<br>$n_t^{\text{recovered}} = \gamma_t I_{t-t_{\text{recovery}}} + \gamma_t I'_{t-t_{\text{recovery}}}$ |
| $n_t^{\text{deaths}}$ | The number of deaths on day $t$<br>$n_t^{\text{deaths}} = m_t I_{t-1} + m_t I'_{t-1}$ |
| $i_t^{\text{first time}}$ | The number of first time infections on day $t$. i.e. the number of $S_t \rightarrow I_t$ transitions on day $t$ |

When using the model, the rate of infection, recovery and fatality for each state were assumed to be independent of how many previous infections a host had previously had. The number of susceptible persons at the beginning of the simulation, N, was taken to be the population of the region of interest [9,12]. After all infections, recoveries and deaths for a day, the number of days into the simulation was increased by one, t→t+1 up to t_max_. The simulation was repeated 10,000 times to produce expectation values and standard deviations for the number of individuals classified as reinfections.

By pooling the number of cases in the Infected (two or more times), Recovered (two or more times) and those deceased from the Infected (two or more times) states at the end of the simulation, we calculated an estimate of the number of reinfections that would be expected to occur. This number represents the total population that had passed through the infected (two or more times) state by the end of the simulation.

Unless otherwise stated, the average recovery time used in the simulations was set as 28 days, as this is greater than the median recovery time suggested in the report of the WHO-China joint mission on coronavirus disease 2019 [14].

## Results

### Simulations of United Kingdom infection data suggest a low number of reinfections should have occurred

We initially ran the simulation for data in the United Kingdom over the course of 106 days (from the first recorded case on the 1^st^ of February until the 17^th^ of May 2020 when the data was accessed). Figure 2 shows how the population of each state in the model changed over the course of a typical simulation. The number of susceptible individuals initially remained steady, until day 55, when there was a sharp decline due to the increase in primary infections (Figure 2A). The number of individuals infected just once started to increase steadily after day 40, and continued to do so throughout the simulation until day 92. After the 28-day lag time, the individuals infected once started to recover, resulting in an increase in the Recovered (once) state through to the end of the simulation (Figure 2B). As the number of recovered individuals started to increase, so did the number of people infected for a second time. The number of people recovered for the second time started to increase after the 28 day recovery lag time (Figure 2C). The number of deaths started to rise from day 55 onwards, and fatalities continued to increase through to the end of the simulation (Figure 2D). In the United Kingdom, the number of expected reinfections was calculated to be 43±7, which makes up 0.018% of the total infections (Table 2). The first reinfection for the UK was on day 82±5, corresponding to the 22^nd^ of April 2020.

**Figure 2.**
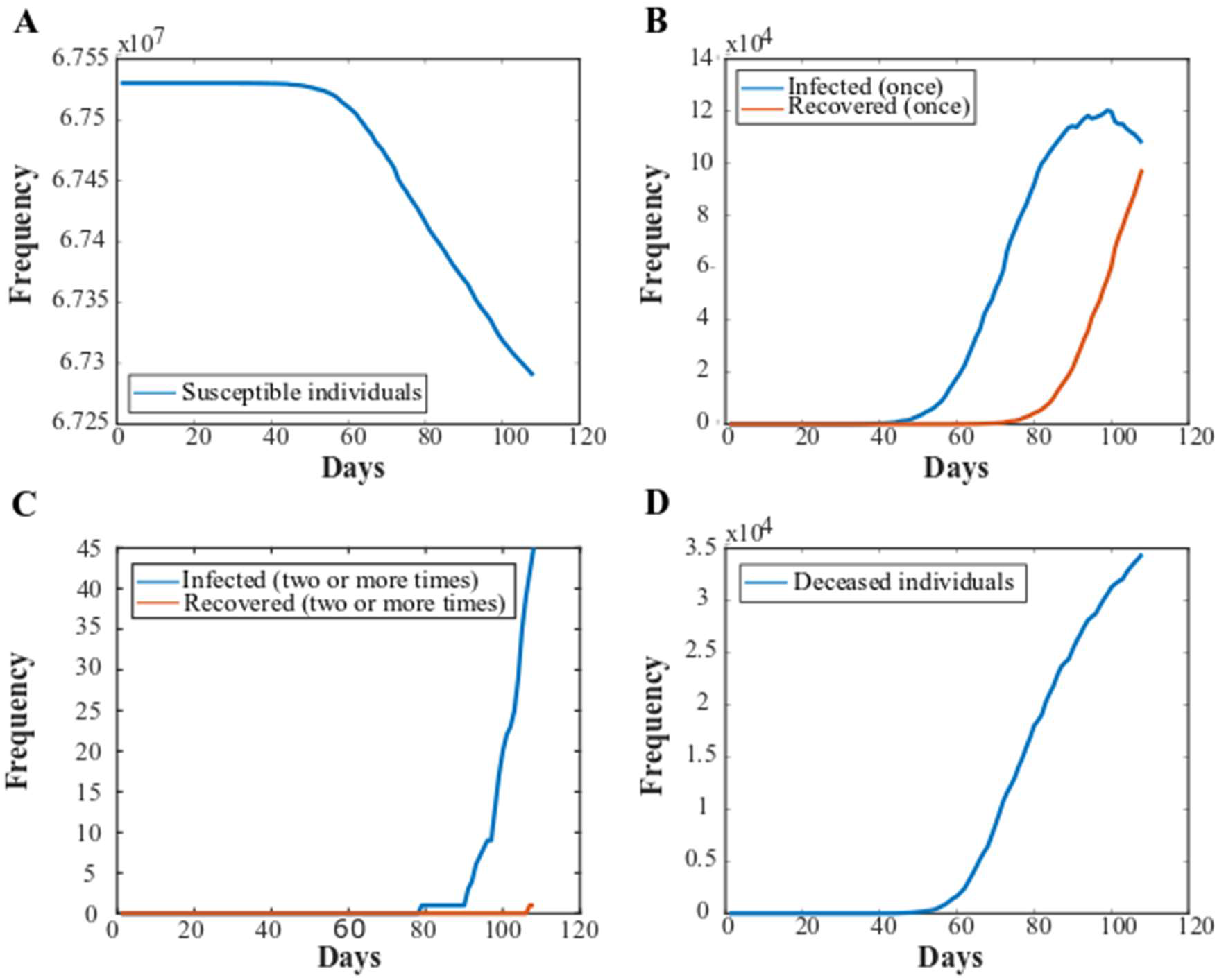
Plots of the populations of each state in the model over the course of a typical simulation, using infection data from the United Kingdom. A) An example plot of the susceptible population in the model over the course of 106 days (from the first recorded case on the 1^st^ of February until the 17^th^ of May 2020 when the data was accessed). B) An example plot of the populations that are infected for the first time or recovered from a single infection. C) An example plot of the populations of the simulation that have been reinfected and recovered from infection twice. D) An example plot of the number of deceased individuals through the course of the simulation.

### Simulations of infection data in other regions show a similar trend

The simulations were repeated with data from Australia, Italy, New York City, Singapore, Switzerland and the United States of America (USA). The mean number of expected reinfections in each region or country for the 10,000 simulations that were run are shown in Table 2. In all cases, with the exception of Australia, our model predicts that reinfection cases should occur.

**Table 2.** The number of predicted reinfections and their standard deviation in different locations worldwide as predicted from the model. Unless otherwise stated, these figures represent simulations using the total number of infections of each region and are modelled without the data on number of recoveries. S.D. standard deviation.

| Region | Mean number of reinfections | Standard deviation of number of reinfections | Total number of infections | Reinfections as a % of total infections |
| --- | --- | --- | --- | --- |
| Australia | 0.1 | 0.3 | 7036 | 0.0014 |
| Germany | 20 | 5 | 174355 | 0.011 |
| Germany (with recovery data) | 79 | 9 | 174355 | 0.05 |
| Italy | 63 | 8.0 | 224760 | 0.028 |
| New York City | 209 | 15 | 189027 | 0.11 |
| New York City (hospitalisations) | 7 | 3 | 48462 | 0.004 |
| Singapore | 4 | 2 | 27356 | 0.014 |
| Switzerland | 4 | 2 | 30587 | 0.013 |
| United Kingdom | 43 | 7 | 240161 | 0.018 |
| United States of America | 402 | 20 | 1467884 | 0.027 |

### Comparison of infection and hospitalisation data in New York City

Next, we repeated our simulation for New York City, with the total number of infections replaced by the number of hospitalisations. When we ran the simulation with an input of the total number of infections, the number of secondary infections continued to increase throughout the simulation, when the numbers appear to start to peak (Figure 3A). This was followed by an increase in the number of secondary recoveries after the 28-day recovery lag time. In comparison, the hospitalisation data for New York showed no secondary recoveries as the reinfections occurred later into the simulation (Figure 3B). The total number of predicted reinfections from the New York hospitalised data was 12±4 (Table 2).

**Figure 3.**
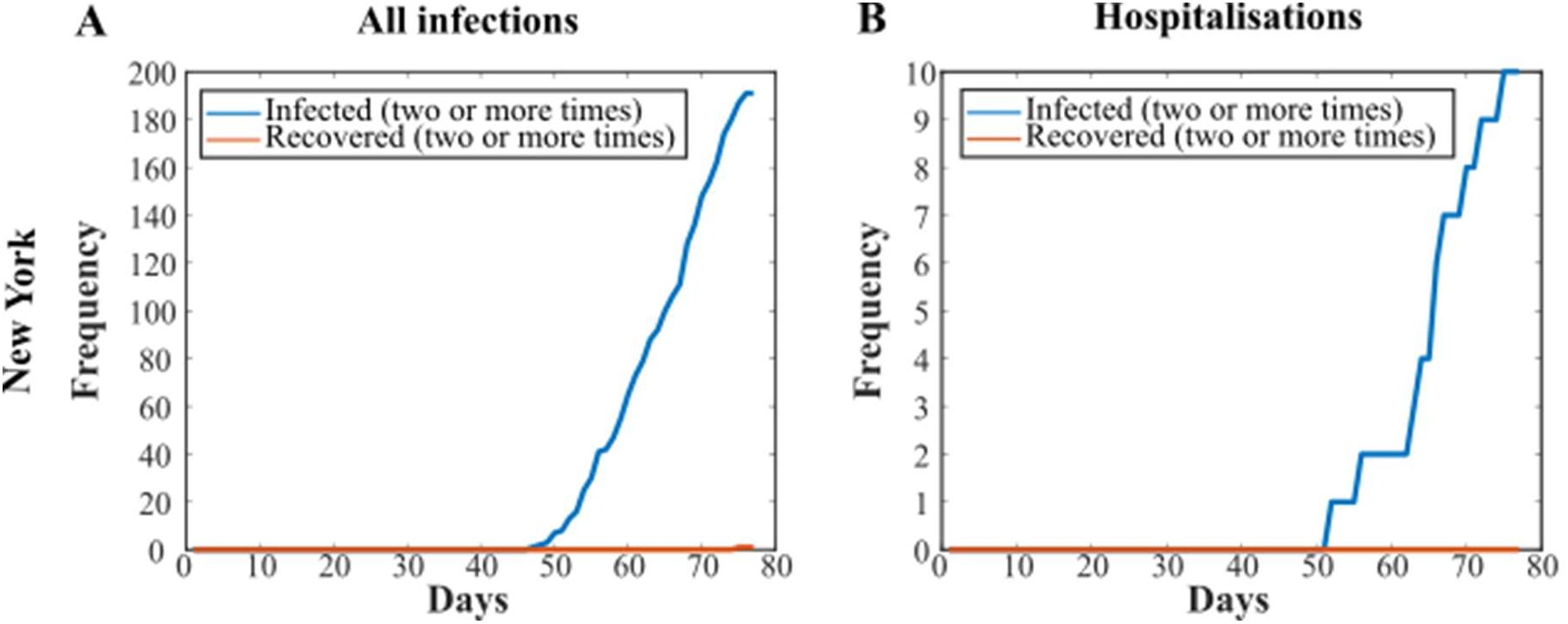
Comparison of total infections vs. hospitalisation data in New York City. Plots of the Infected (two or more times) and Recovered (two or more times) states for A) New York using all infection data B) New York using only the hospitalisations data.

### Inclusion of recovery data suggests that predicted reinfections are underestimated

Recovery data was sparse or unavailable for most regions, likely due to lack of follow-up testing. There was recovery available from Germany and we therefore compared the results of our simulation for Germany with and without the recovery data as an input. The models used a 28-day lag before recoveries started, meaning very few secondary recoveries took place (Figure 4A and Figure 4B). There were 73 more reinfections with the reinfection data than with the modelled data (Table 2).

**Figure 4.**
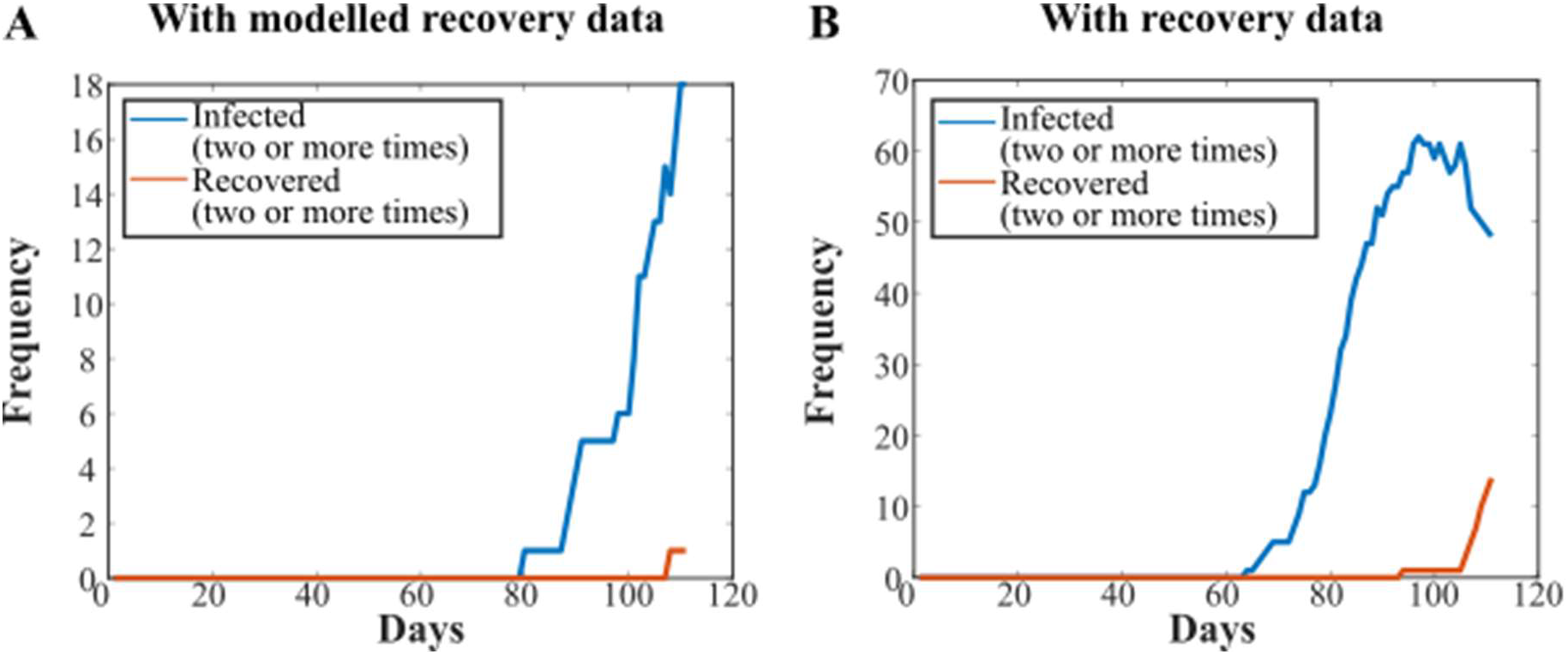
Use of actual recovery data from Germany suggests that the number of recovered individuals, and hence reinfections, are underestimated in our model. Plots of Infected (two or more times) and Recovered (two or more times) states with A) modelled recovery data and B) actual recovery

The 28-day lag time used for the modelled recovery data ensured that we underestimated the recovery rate, and so also the rate of reinfection. To investigate a more life-like recovery rate, the United Kingdom simulations were repeated again using the modelled recovery data, while shortening the lag time for recovery. As expected, we found that the rate of reinfection increased as the lag time was decreased from 28 days through to 7 days, as there was a larger population that recovered from a primary infection. With a 7-day lag time, the number of people in the infected (2 or more times) state peaked at day 101 of the simulation (Figure S1). The total number of people reinfected throughout the simulation increased with a decreasing lag time, with 43±7, 83±9, 139± 12 and 209±14 reinfections for 28-day, 21-day, 14-day and 7-day recovery lag times respectively.

## Discussion

In this work we have presented a modelling strategy used to determine whether SARS-CoV-2 reinfections can occur. We modelled actual infection and fatality data from different regions around the world and found that all regions investigated, with the exception of Australia, should have recorded cases of reinfections if primary infection with SARS-CoV-2 did not provide some level of immunity. The actual number of cases of reinfection that have been reported in any of these regions or countries to date is zero, suggesting that worldwide, primary SARS-CoV-2 infection provides short term immunity.

In Australia, the number of confirmed SARS-CoV-2 infections at the time the data was accessed was relatively low [9], possibly due to early social distancing measures, the closing of international borders and mass testing and tracing measures. The number of modelled reinfections (0.1±0.3; Table 2) reflects this, and so even without immunity from infection no reinfections would be expected to occur. Similarly, in Switzerland and Singapore, very low numbers of reinfections were predicted by the model (6.2±2.5 and 6±2 respectively; Table 2). It is possible that these very low numbers of reinfection cases could have been missed due to misdiagnosis or lack of follow up testing. We therefore applied our model to data from Germany [9], Italy [9], New York City [11,12] and the USA as a whole [9], which have recorded far higher numbers of SARS-CoV-2 infections (174355, 224760, 189027 and 1467884 respectively, when the data was accessed). The number of reinfection cases predicted for these countries was 30±6, 89±9, 335±18 and 635±25 for Germany, Italy, New York and the USA respectively (Table 2). We conclude that it is therefore very unlikely that all of these predicted cases, if true, were missed due to misdiagnosis or lack of testing.

We also found that rehospitalisation cases should have been seen amongst hospitalised cases in New York City - it is unlikely that these cases would be missed as people are processed and tested on admission into hospital. To date, however, no reinfections have verifiably been recorded anywhere in the world. A report from South Korea suggested that 116 patients recovered from COVID-19 had tested positive by RT-PCR for the virus again [18], however, this has since been explained as the ‘false-positive’ detection of remnants of viral RNA rather than reactivation or reinfection. The lack of documented reinfections suggests that short-term immunity to the virus is produced by an initial infection, however, our model cannot predict whether this immunity will last over longer time scales.

Our results are supported by a number of animal challenge studies which also show immunity to SARS-CoV-2 can be conferred. A study in rhesus macaques showed that, following initial viral clearance, the monkeys showed a reduction in their median viral load in comparison with primary infection when rechallenged with SARS-CoV-2 [19]. Similarly, Ryan et al. demonstrated that rechallenged ferrets were fully protected from acute lung pathology [20]. An adenovirus-vector vaccine tested on rhesus macaques elicited a humoral and cellular response that, on challenge with the virus, proved to significantly reduce the viral load in bronchoalveolar lavage fluid and respiratory tract tissue [21]. However, a longitudinal study by Seow et al. [22] showed that the immunity conferred against SARS-CoV-2 may only be short-term. Our model proposes that reinfection cases should have already started to appear by April 2020, suggesting a possible lower limit for immunity duration.

A report from the WHO-China joint mission on Coronavirus disease estimated the recovery time for SARS-CoV-2 infection as 2 weeks for mild cases and 3-6 weeks for severe or critical disease [14]; based on this we used a long (28 day) recovery lag time in the modelled data. Comparison with real-world recovery data from Germany suggested that the actual recovery time may be significantly shorter, giving rise to an underestimation of the reinfection rate in our modelled data. This was supported by an increase in the number of predicted reinfections in the United Kingdom simulations when we used a shorter recovery lag time of 7, 14 or 21 days. In addition, there were no allowances in our model for transmission being localised to regions smaller than a nation or city; the daily infection data was likely to be only a fraction of the total number of infections due to asymptomatic or mild infections not being recorded, and infections were recorded on the date of testing, not the actual date of infection. We also note that significant differences in testing, reporting and shielding of the vulnerable exist between the different regions in this study and that a large number of SARS-CoV-2 cases were missed in every region of interest (for example, in Geneva, unreported cases were estimated to be 11.6 infections per reported infection from April 6 to May 9 [17]). In every region, we expect that the impact on our simulation would be to underestimate the number of reinfections. Taken together, this suggests that the actual reinfection rate would be significantly higher than that predicted by our model if there was no immunity conferred by prior infection.

Our model has a number of limitations, including the lack of modelling of any social structure, the fact that individuals who have been infected may change their shielding behaviours, differing recovery times from person to person, and missing information regarding immigration into and out of regions of interest. In spite of this, the results documented here provide strong evidence, based on real data, to suggest that that there is at least short-term immunity conferred by an initial infection of SARS-CoV-2. This has implications for serological testing strategies, lockdown easing timescales and vaccine development. Our modelling strategy can also be extended to understand the reinfection dynamics of future pandemics.

## Data Availability

The data that support the findings of this study are openly available at https://ourworldindata.org/coronavirus-country-by-country, reference number 19; https://www1.nyc.gov/site/doh/covid/covid-19-data.page, reference number 22; https://www.bag.admin.ch/bag/en/home/krankheiten/ausbrueche-epidemien-pandemien/aktuelle-ausbrueche-epidemien/novel-cov/situation-schweiz-und-international.html#-1199962081, reference number 23, and https://tradingeconomics.com/germany/coronavirus-recovered, reference number 24.

https://www1.nyc.gov/site/doh/covid/covid-19-data.page

https://www.bag.admin.ch/bag/en/home/krankheiten/ausbrueche-epidemien-pandemien/aktuelle-ausbrueche-epidemien/novel-cov/situation-schweiz-und-international.html#-1199962081

https://tradingeconomics.com/germany/coronavirus-recovered

## Acknowledgements

We thank Dr. Barak Gilboa for critical reading of the manuscript. This work was supported by Royal Society Dorothy Hodgkin Research Fellowship DKR00620 and Research Grant for Research Fellows RGF\R1\180054 (to N.C.R.).

